# Predictors of poor glycemic control among adults attending a peri-urban diabetic clinic in Wakiso district, Uganda: A cross-sectional study using modified Poisson regression analysis

**DOI:** 10.64898/2026.06.02.26354687

**Authors:** Larissa Ngazet Yangbalet Kezza, Ronald Kooko, David Musoke, Elizeus Rutebemberwa, Olivia Nakisita, Marius Wilfried Wanikomane Dandy, Pierre Somse

## Abstract

**Background:** Poor glycemic control, a contributor to the development of diabetes related complications among patients with diabetes mellitus, remains a public health challenge in low-and middle-income countries. In Uganda, limited evidence exists on predictors of poor glycemic control among diabetic patients attending peri-urban primary healthcare facilities. The study assessed predictors of poor glycemic control among adults attending the diabetic clinic at Kasangati Health Centre IV in Wakiso district.

**Methods:** We conducted cross-sectional study among 283 diabetic patients attending Kasangati Health Centre IV between March and April 2025. Data were collected using interviewer-administered structured questionnaires and data abstraction tools. Poor glycemic control was defined as glycated hemoglobin (HbA1c) levels ≥7%. Modified Poisson regression with robust standard errors was used to determine factors associated with poor glycemic control. Adjusted prevalence ratios (aPRs) with 95% confidence intervals (CIs) were reported.

**Results:** Overall, 67.8% of the participants had poor glycemic control. Poor glycemic control was significantly associated with older age, low income status (aPR: 1.4, 95%CI: 1.24–1.58), use of multiple anti-diabetic medications, non-adherence to regular follow-up (aPR: 1.5, 95%CI: 1.33–1.65), medication side effects (aPR: 1.2, 95%CI: 1.01–1.32), physical inactivity (aPR: 1.1, 95%CI: 1.05–1.21), non-adherence to recommended dietary plans (aPR: 1.1, 95%CI: 1.02–1.22), perceived treatment regimen complexity (aPR: 1.2, 95%CI: 1.12–1.34), stress (aPR: 1.1, 95%CI: 1.08–1.20), lack of peer support groups (aPR: 1.2, 95%CI: 1.08–1.23), and high costs of accessing care (aPR: 1.2, 95%CI: 1.17–1.33).

**Conclusion:** Almost two-thirds of the diabetic patients suffered from poor glycemic control which was determined by various socio-economic, behavioral, clinical and health system factors. Enhancing adherence counseling, encouraging healthy lifestyles, adopting age-based supportive healthcare approaches, better psychosocial support and reduction of cost barriers in accessing diabetic healthcare could improve the glycemic status of diabetic patients in peri-urban primary healthcare settings.

## Introduction

Diabetes Mellitus (DM) is one of the major non-communicable diseases and is defined as a condition where hyperglycemia persists due to impairment in insulin production, insulin resistance, or both [1]. The global prevalence of diabetes is increasing at a fast pace, with 537 million adult patients (20-79 years old) being reported in 2021, and projections indicate an increase to 783 million by 2045 [2]. More than three-quarters of people living with diabetes reside in low-and middle-income countries where healthcare systems often face challenges related to inadequate financing, limited infrastructure, and shortages of trained healthcare workers [3, 4].

In sub-Saharan Africa, there has been a noticeable growth in the prevalence of DM with the number of cases expected to increase from 24 million in 2021 to 55 million in 2045, an approximately 129% increase, which is the highest rate of increment seen worldwide [1, 2, 5]. In Uganda, the burden of diabetes mellitus has increased over the past decade, with national prevalence rising from 1.5% in 2014 to approximately 3.3% in 2023, and urban prevalence increasing from 2.3% to 3.6% over the same period [6]. This growing burden has been attributed to rapid urbanization, unhealthy dietary habits, physical inactivity, and increasing adoption of sedentary lifestyles, particularly in urban and peri-urban settings [7].

Glycemic control, measured in terms of glycated hemoglobin (HbA1c), is an important indicator of diabetes management. Optimal glycemic control with HbA1c values less than 7% has been linked to a lower risk of diabetes complications and improved quality of life [8]. Nevertheless, optimal glycemic control is not easy to achieve for many patients in resource-limited settings due to multiple interacting factors including low income, medication burden, poor compliance with dietary advice, physical inactivity, stress, lack of social support, and high healthcare costs [9, 10]. Previous studies in Uganda and other low-resource settings have also shown that medicine shortage, poor patient education, and weak health systems negatively influence diabetes outcomes [11, 12].

Wakiso district is one of the fastest urbanizing districts in Uganda, experiencing an increasing burden of non-communicable diseases, including diabetes mellitus. Kasangati Health Centre IV, which is a high-volume peri-urban primary healthcare facility, offers diabetic care services to the rising number of patients seeking medical attention. Despite the availability of diabetic care services, anecdotal reports and facility observations suggest persistent challenges in achieving optimal glycemic control among patients attending the clinic. However, limited information about predictors of poor glycemic control among patients seeking diabetic care services in peri-urban primary healthcare facilities in Uganda. This study therefore, assessed predictors of poor glycemic control among adults accessing diabetic care services at Kasangati Health Centre IV in Wakiso district using modified Poisson regression analysis. Findings from this study may inform targeted interventions aimed at improving diabetes management and reducing diabetes-related complications in similar peri-urban primary healthcare settings

## Materials and Methods

### Ethical considerations

This study was reviewed and approved by the Makerere University, School of Public Health Research and Ethics Committee under reference number; MakSPH-REC_502. Permission to conduct the study was also obtained from the administration of Kasangati Health Centre IV. Before recruitment, the purpose and procedures of the study were explained to potential participants, and participation was voluntary. Written informed consent was obtained from all participants prior to enrolment into the study. Confidentiality was maintained throughout the study, and all information collected was used solely for research purposes.

### Study design and setting

This was a facility-based cross-sectional study involving quantitative data collection approach at Kasangati Health Centre IV in Wakiso district, central Uganda. Kasangati Health Centre IV is a peri-urban public health facility located in Nangabo sub-county, approximately 16 kilometers from Kampala city. The health facility provides a wide range of services including outpatient and inpatient care, maternal and child health services, laboratory services, and management of chronic non-communicable diseases including diabetes mellitus. The diabetic clinic at Kasangati Health Centre IV provides services such as patient consultation, routine follow-up, blood glucose monitoring, lifestyle counselling, screening for diabetes-related complications, and treatment for diabetic patients. The facility serves a rapidly growing peri-urban population with an increasing burden of diabetes mellitus and other non-communicable diseases.

### Inclusion criteria

The study included adult patients aged 18 years and above diagnosed with diabetes mellitus and attending the diabetic clinic at Kasangati Health Centre IV during the study period. Participants who had attended the diabetic clinic for at least six months were included to ensure that they had adequate exposure to diabetes care services at the facility and could provide reliable information regarding glycemic control and diabetes management practices.

### Exclusion criteria

The study excluded severely ill patients who were unable to participate in the interviews and pregnant women with gestational diabetes because their management approaches and glycemic outcomes differ from those of the general adult diabetic population.

### 3.5 Sample size determination

The sample size was determined using Taro Yamane formula was used as expressed below,

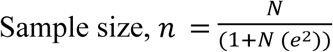

Where n is the minimum sample size, N is the population under study (number of DM patients attending the diabetic clinic at Kasanganti health Centre IV) which estimated at 700 and e is the error i.e., 5%.

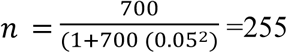

Considering a non-response rate of 10%

Final sample size= Effective sample size/ (1-non response rate anticipated) n= 255 / (1-10%) ∼ **283** DM patients

### Sampling procedure

A systematic random sampling technique was used to select study participants attending the diabetic clinic at Kasangati Health Centre IV. The diabetic clinic register was used as the sampling frame to identify eligible participants seeking diabetes care during the study period. The sampling interval (k) was determined by dividing the total number of registered diabetic patients (N = 700) by the calculated sample size (n = 283), resulting in a sampling interval of k ≈ 3. Using a random number generator, a starting point between 1 and 3 was selected to identify the first participant. Thereafter, every 3rd patient on the register was selected until the desired sample size was attained. This approach ensured an even distribution of participants throughout the sampling frame, minimizing selection bias.

### Study variables

The dependent variable for the study was glycemic control among diabetic patients, assessed using glycated hemoglobin (HbA1c) levels. Poor glycemic control was defined as HbA1c levels ≥7%, while good glycemic control was defined as HbA1c levels <7%. The independent variables included socio-demographic factors (age, sex, marital status, level of education, occupation, income level, and area of residence), clinical factors (duration of treatment, number of medications, comorbidities, anti-diabetic treatment regimen, and medication side effects), behavioral factors (physical exercise, dietary practices, blood sugar monitoring, and stress), and health system factors (support groups, costs of accessing care, and receipt of diabetes care package).

### Data collection

Trained research assistants with support from the principal investigator collected data over a period of one month from 6^th^ February 2025 to 6^th^ March 2025 using pre-tested interviewer-administered structured questionnaires. The data collection tools were adapted from literature and previously validated tools used in similar studies on diabetes management and glycemic control (Faisal et al., 2022). The questionnaire collected information on socio-demographic characteristics, clinical factors, individual factors, patient practices, and health system-related factors associated with glycemic control among diabetic patients. Clinical information including HbA1c levels, anti-diabetic medications, duration of treatment, and comorbidities were obtained from patient medical records and clinic registers using a data abstraction tool.

### Training of research assistants

The principal investigator selected the research assistants, trained them for four days on questionnaire administration, data collection, and ethical considerations; they were also trained in effective communication skills, showing empathy and never being judgmental. The research assistants were aged between 20 and 30 years and fluent in the local language and attained at least certificate qualification with some holding diploma and bachelor’s degree in health-related or social science field. This helped to ensure free, open communication for the participants and reliable administration of the questionnaire.

### Quality control measures

The data collection tools were translated into Luganda and back-translated into English to ensure consistency in meaning. Pre-testing of the tools was conducted among 15 diabetic patients attending Goma Health Centre IV a peri-urban health facility in Mukono district with similar characteristics to Kasangati Health Centre IV. Internal consistency reliability was assessed using Cronbach’s alpha, yielding a coefficient of ≥0.86 across key domains, indicating acceptable reliability. Feedback from pretesting informed refinement of question wording and sequencing. Fieldwork was overseen by the principal investigator to ensure completeness and accuracy of data collected.

### Data analysis

Data analysis was performed using Statistical Package for Social Sciences (SPSS) version 26. Descriptive statistics were used to summarize participants’ characteristics, with categorical variables presented as frequencies and percentages. Prior to multivariable modeling, multicollinearity among independent variables was assessed using the variance inflation factor (VIF), and all variables had VIF values below 5, indicating absence of significant multicollinearity. Bivariate analysis was conducted using modified Poisson regression to determine factors associated with poor glycemic control among diabetic patients. Variables with p-values less than 0.2 at bivariate analysis were considered for multivariable analysis to avoid excluding potential predictors. Modified Poisson regression with robust standard errors was used to estimate adjusted prevalence ratios (aPRs) and 95% confidence intervals (CI) because the outcome was common (>10%). Model fit was assessed using the Omnibus test (X^2^ = 34.3, df = 21, p< 0.001), indicating an excellent model fit. Adjusted prevalence ratios (aPRs) with 95% CI were reported, and statistical significance was considered at p<0.05.

## Results

### Socio-demographic characteristics of the participants

A total of 283 adults attending the diabetic clinic at Kasangati Health Centre IV were included in this study. The mean age of the participants was 40.2 ± 13.2 years, while the median age was 41 years (IQR: 29–50 years). Nearly half of the participants (49.8%) were aged 40–64 years, whereas 44.2% were aged <40 years and only 6.0% were aged ≥65 years. More than half of the respondents were female (59.0%), married (50.2%), and residing in rural areas (53.7%). Most participants had low income levels (68.6%) and were peasants (60.8%). Regarding education level, 39.6% had attained secondary education while 15.5% had no formal education. Catholics constituted the largest religious group (34.6%), followed by Anglicans (29.7%) and Pentecostals (25.1%) (Table 1).

**Table 1:** Socio-demographic characteristics of diabetes mellitus patients at Kasangati HCIV.

| Variable | Frequency | Percentage |
| --- | --- | --- |
| **Age** |  |  |
| Age (Mean ± SD) | | 40.2±13.2 |
| Age (Medium, IQR) |  | 41 (29,50) |
| <40 | 125 | 44.2 |
| 40-64 | 141 | 49.8 |
| $\geq 65$ | 17 | 6.0 |
| **Marital status** |  |  |
| Single | 99 | 35.0 |
| Married | 142 | 50.2 |
| Divorced | 25 | 8.8 |
| Widowed | 17 | 6.0 |
| **Gender** |  |  |
| Male | 116 | 41.0 |
| Female | 167 | 59.0 |
| **Income level** |  |  |
| Low | 194 | 68.6 |
| High | 89 | 31.4 |
| **Occupation** |  |  |
| Unemployed | 21 | 7.4 |
| Peasants | 171 | 60.8 |
| Business | 66 | 23.3 |
| Civil Servants | 24 | 8.5 |
| **Level of education** |  |  |
| None | 44 | 15.5 |
| Primary | 106 | 37.5 |
| Secondary | 112 | 39.6 |
| Tertiary | 21 | 7.4 |
| **Area of residence** |  |  |
| Rural | 152 | 53.7 |
| Urban | 131 | 46.3 |
| **Religion** |  |  |
| Anglican | 84 | 29.7 |
| Catholic | 98 | 34.6 |
| Muslim | 30 | 10.6 |
| Pentecostal | 71 | 25.1 |

### Glycemic control among participants

Overall, 67.8% of the participants had poor glycemic control, while only 32.2% had good glycemic control based on HbA1c measurements (Figure 1).

**Figure 1:**
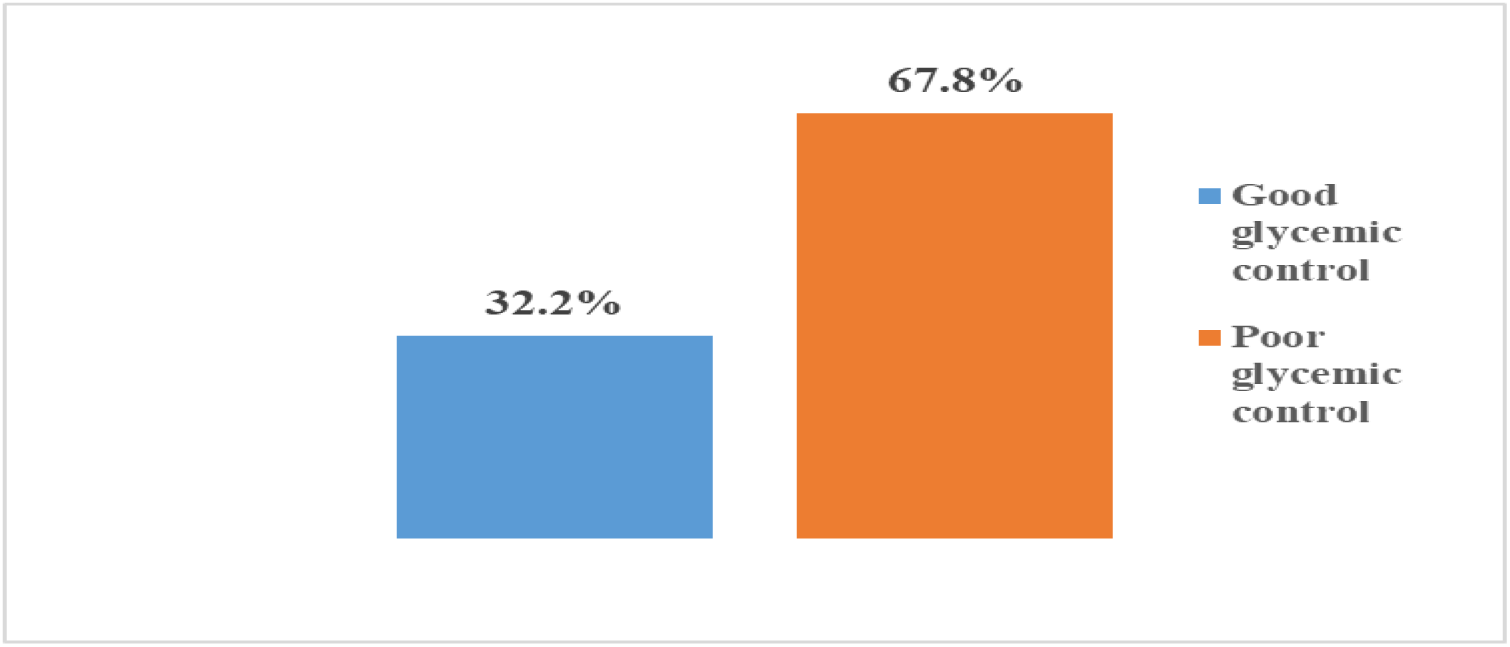
Glycemic levels (HbAIC) among DM patients at Kasangati HCIV

### Bivariate analysis of factors associated with poor glycemic control

#### Socio-demographic factors associated with poor glycemic control

Participants aged 40–64 years had a significantly higher prevalence of poor glycemic control (74.5%) compared to those aged <40 years (60.8%) and were 1.1 times more likely to have poor glycemic control (cPR = 1.1, 95% CI: 1.01–1.26). Similarly, participants aged ≥65 years had a high prevalence of poor glycemic control (67.8%), although the association was not statistically significant.

Poor glycemic control was markedly higher among participants with low income levels (90.2%) compared to those with high income levels (19.1%). Participants with low income were 1.6 times more likely to experience poor glycemic control than their counterparts with high income (cPR = 1.6, 95% CI: 1.49–1.72). Education level was also significantly associated with glycemic control. Participants who had attained secondary education and tertiary education were less likely to experience poor glycemic control compared to those with no formal education. Similarly, rural residents had a substantially higher prevalence of poor glycemic control (84.9%) compared to urban residents (48.1%), and were 1.3 times more likely to have poor glycemic control (cPR = 1.3, 95% CI: 1.17–1.33). (Table 2).

**Table 2.**
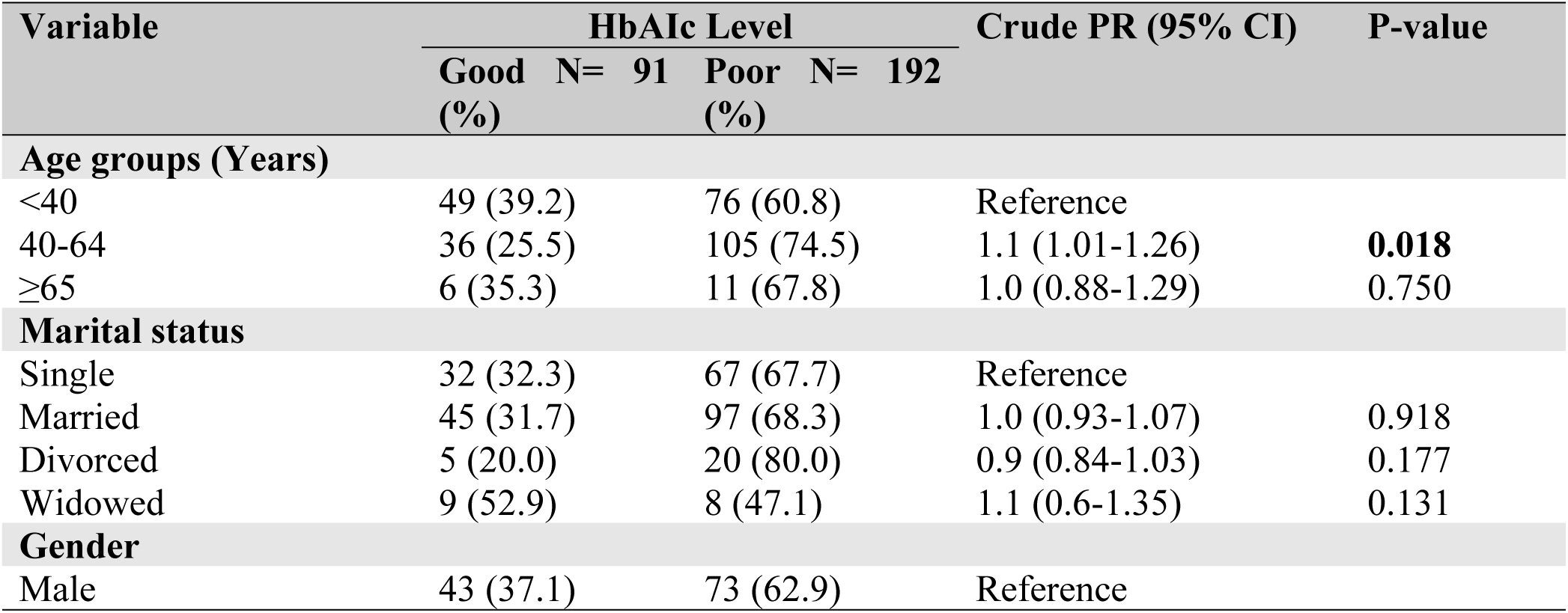
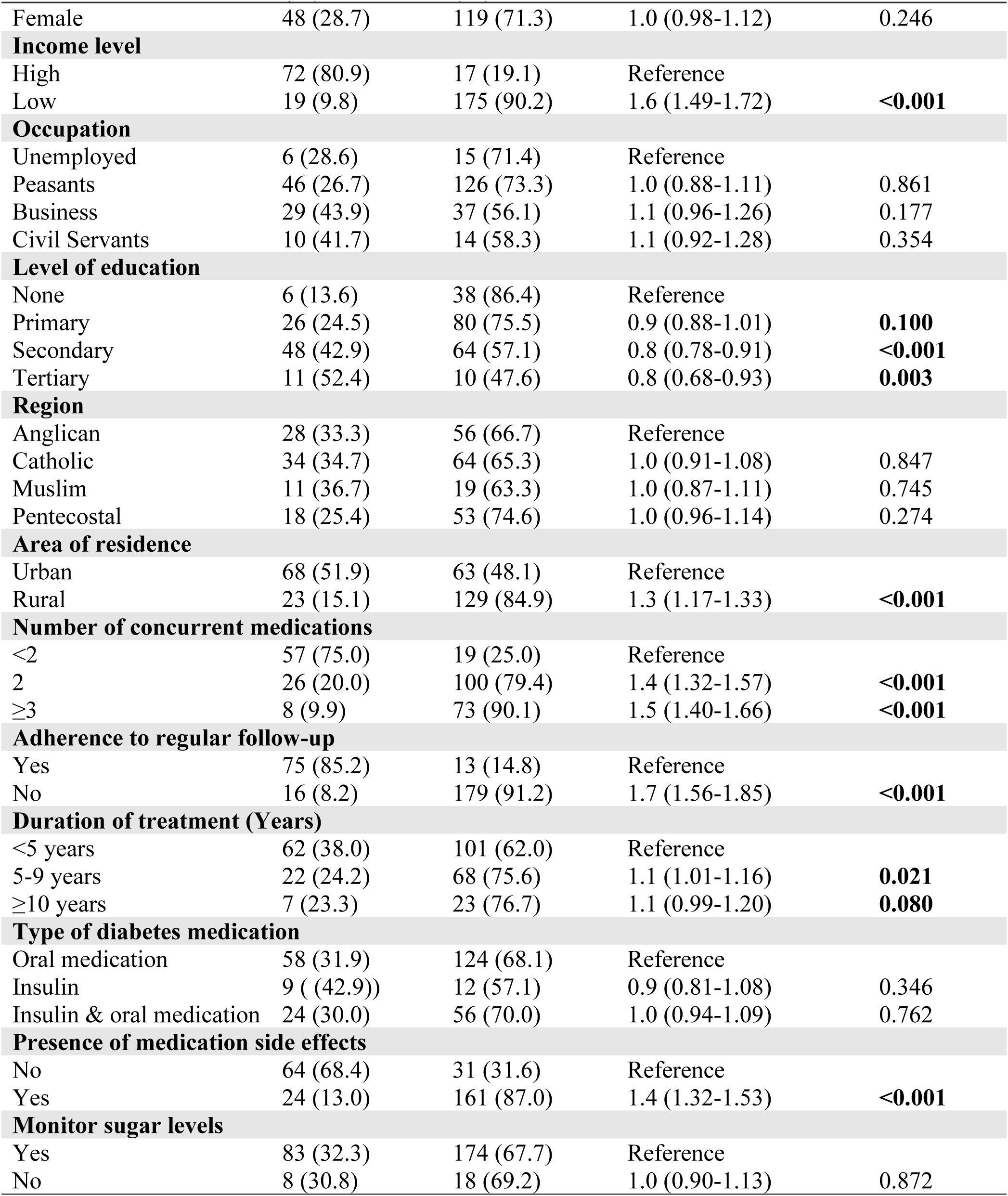
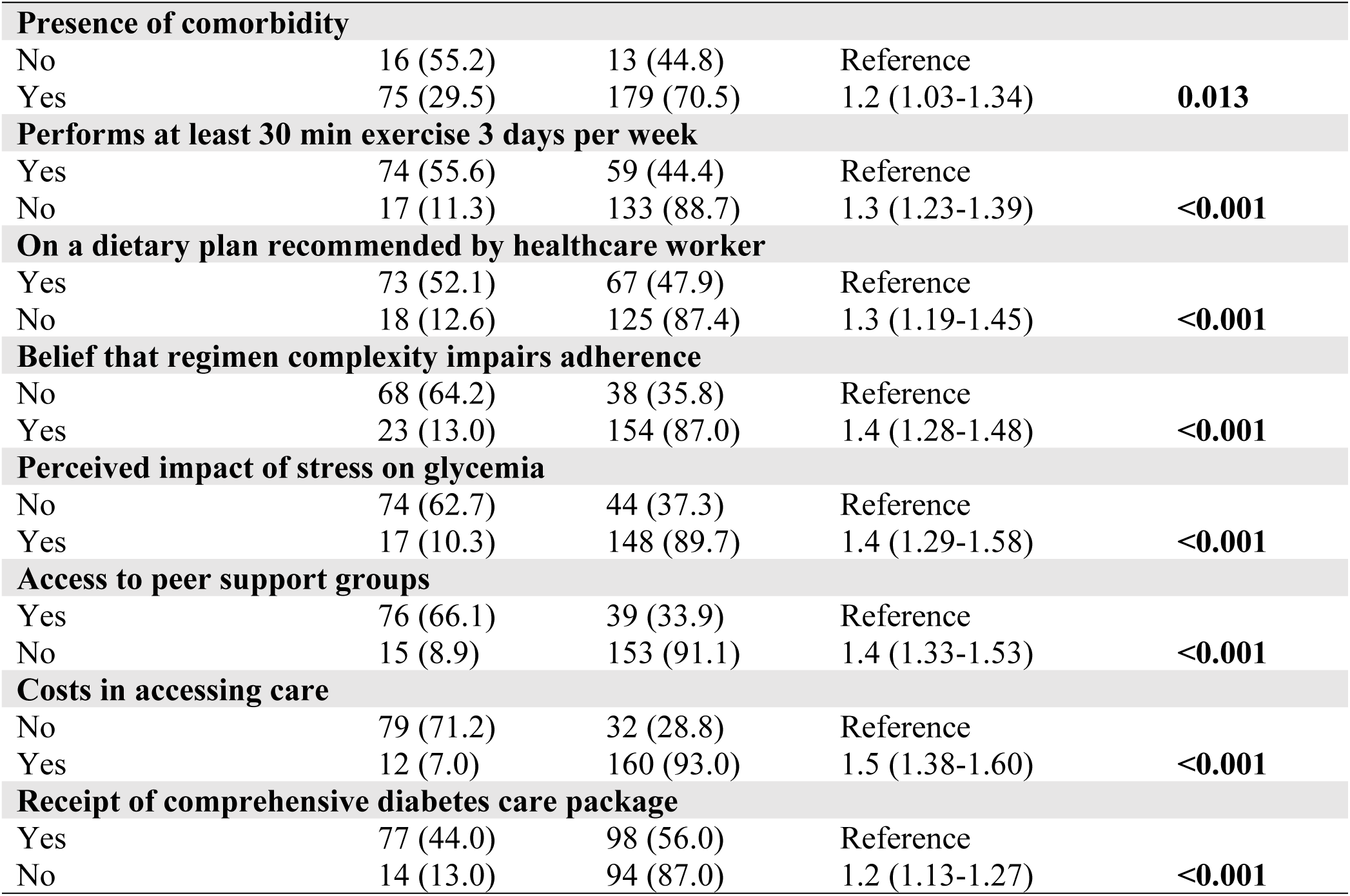
Bivariate analysis of factors associated with poor glycemic control. N=283.

#### Clinical and behavioral factors associated with poor glycemic control

Participants taking two concurrent medications and those taking three or more medications had significantly higher prevalence of poor glycemic control compared to those taking fewer than two medications. Those taking ≥3 medications were 1.5 times more likely to have poor glycemic control (cPR = 1.5, 95% CI: 1.40–1.66).

Non-adherence to regular clinic follow-up was strongly associated with poor glycemic control. Participants who did not adhere to regular follow-up had a prevalence of poor glycemic control of 91.2% compared to 14.8% among those who adhered to follow-up schedules. These participants were 1.7 times more likely to experience poor glycemic control (cPR = 1.7, 95% CI: 1.56–1.85). Participants with medication side effects had significantly higher prevalence of poor glycemic control (87.0%) compared to those without side effects (31.6%). Similarly, participants with comorbidities were more likely to have poor glycemic control than those without comorbidities. Lifestyle-related factors were also significantly associated with poor glycemic control. Participants who did not perform at least 30 minutes of exercise three days per week had significantly higher prevalence of poor glycemic control (88.7%) compared to those who exercised regularly (44.4%). Similarly, participants who were not on a dietary plan recommended by healthcare workers had higher prevalence of poor glycemic control (87.4%) than those following recommended dietary plans (47.9%). Psychosocial and health-system-related factors including perceived regimen complexity, stress affecting glycemia, lack of peer support groups, costs in accessing care, and failure to receive a comprehensive diabetes care package were all significantly associated with poor glycemic control at bivariate analysis (Table 2).

### Multivariable analysis of factors associated with poor glycemic control

All variables with p-values less than 0.20 at bivariate analysis were included in the multivariable modified Poisson regression model. The final model depicts the best fit for this analysis. Variables with p-values <0.05 in the adjusted model were considered independently associated with poor glycemic control.

#### Factors independently associated with poor glycemic control

The factors that remained significantly associated with poor glycemic control at multivariable analysis included age 40–64 years (aPR = 1.1, 95% CI: 1.02–1.19), age ≥65 years (aPR = 1.1, 95% CI: 1.00–1.20), low income level (aPR = 1.4, 95% CI: 1.24–1.58), taking two concurrent medications (aPR = 1.1, 95% CI: 1.03–1.20), taking three or more concurrent medications (aPR = 1.2, 95% CI: 1.08–1.31), non-adherence to regular follow-up schedules (aPR = 1.5, 95% CI: 1.33–1.65), presence of medication side effects (aPR = 1.2, 95% CI: 1.01–1.32), lack of regular physical exercise (aPR = 1.1, 95% CI: 1.05–1.21), failure to follow a recommended dietary plan (aPR = 1.1, 95% CI: 1.02–1.22), belief that treatment regimen complexity impairs adherence (aPR = 1.2, 95% CI: 1.12–1.34), perceived impact of stress on glycemia (aPR = 1.1, 95% CI: 1.08–1.20), lack of access to peer support groups (aPR = 1.2, 95% CI: 1.08–1.32), and costs involved in accessing diabetes care (aPR = 1.2, 95% CI: 1.17–1.33) (Table 3).

**Table 3.**
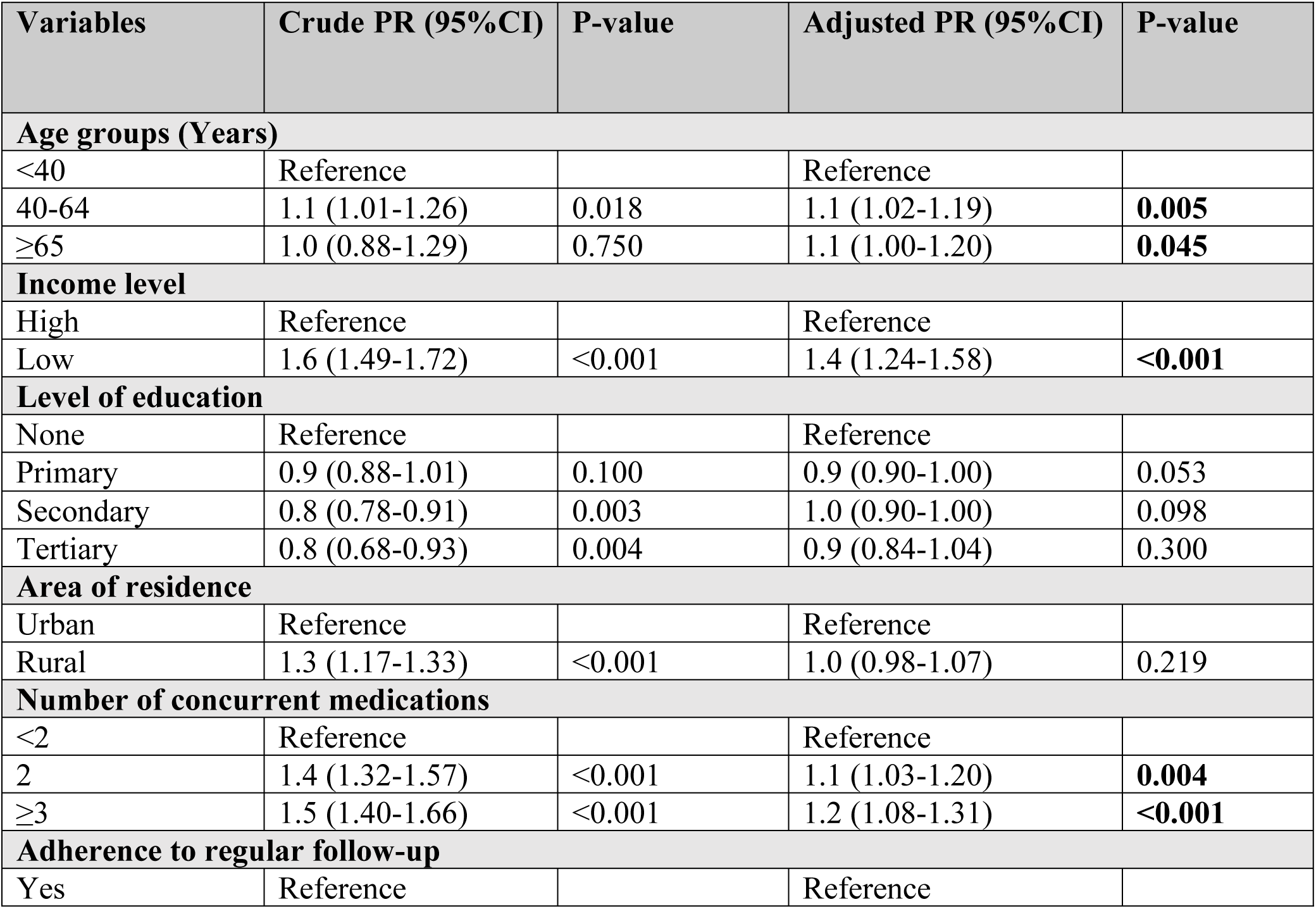
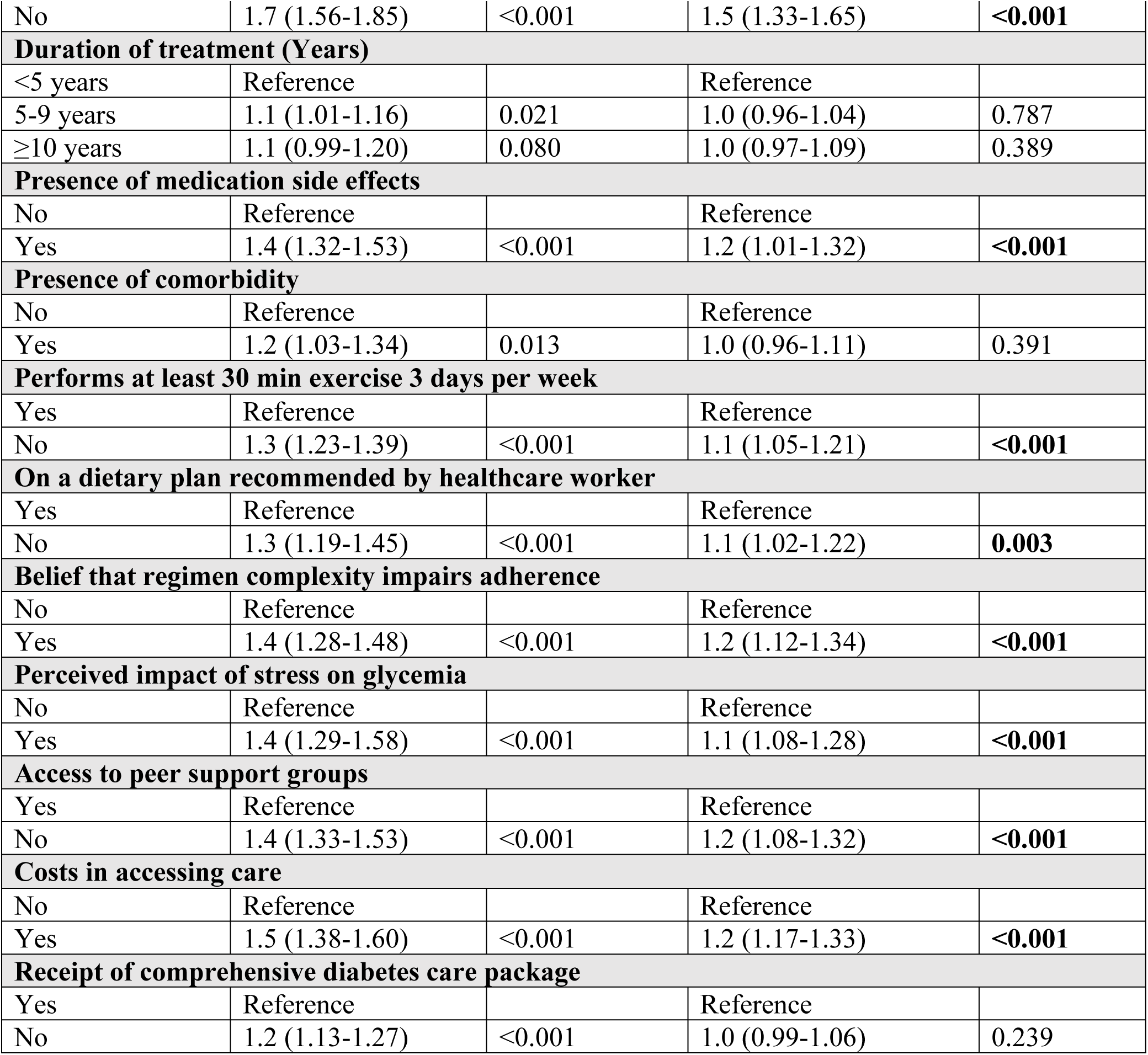
Multivariable analysis of factors associated with Poor glycemic control. N=283

| Variables | Crude PR (95%CI) | P-value | Adjusted PR (95%CI) | P-value |
| --- | --- | --- | --- | --- |
| **Age groups (Years)** |  |  |  |  |
| <40 | Reference |  | Reference |  |
| 40-64 | 1.1 (1.01-1.26) | 0.018 | 1.1 (1.02-1.19) | **0.005** |
| ≥65 | 1.0 (0.88-1.29) | 0.750 | 1.1 (1.00-1.20) | **0.045** |
| **Income level** |  |  |  |  |
| High | Reference |  | Reference |  |
| Low | 1.6 (1.49-1.72) | <0.001 | 1.4 (1.24-1.58) | **<0.001** |
| **Level of education** |  |  |  |  |
| None | Reference |  | Reference |  |
| Primary | 0.9 (0.88-1.01) | 0.100 | 0.9 (0.90-1.00) | 0.053 |
| Secondary | 0.8 (0.78-0.91) | 0.003 | 1.0 (0.90-1.00) | 0.098 |
| Tertiary | 0.8 (0.68-0.93) | 0.004 | 0.9 (0.84-1.04) | 0.300 |
| **Area of residence** |  |  |  |  |
| Urban | Reference |  | Reference |  |
| Rural | 1.3 (1.17-1.33) | <0.001 | 1.0 (0.98-1.07) | 0.219 |
| **Number of concurrent medications** |  |  |  |  |
| <2 | Reference |  | Reference |  |
| 2 | 1.4 (1.32-1.57) | <0.001 | 1.1 (1.03-1.20) | **0.004** |
| ≥3 | 1.5 (1.40-1.66) | <0.001 | 1.2 (1.08-1.31) | **<0.001** |
| **Adherence to regular follow-up** |  |  |  |  |
| Yes | Reference |  | Reference |  |
| No | 1.7 (1.56-1.85) | <0.001 | 1.5 (1.33-1.65) | **<0.001** |
| **Duration of treatment (Years)** |  |  |  |  |
| <5 years | Reference |  | Reference |  |
| 5-9 years | 1.1 (1.01-1.16) | 0.021 | 1.0 (0.96-1.04) | 0.787 |
| ≥10 years | 1.1 (0.99-1.20) | 0.080 | 1.0 (0.97-1.09) | 0.389 |
| **Presence of medication side effects** |  |  |  |  |
| No | Reference |  | Reference |  |
| Yes | 1.4 (1.32-1.53) | <0.001 | 1.2 (1.01-1.32) | **<0.001** |
| **Presence of comorbidity** |  |  |  |  |
| No | Reference |  | Reference |  |
| Yes | 1.2 (1.03-1.34) | 0.013 | 1.0 (0.96-1.11) | 0.391 |
| **Performs at least 30 min exercise 3 days per week** |  |  |  |  |
| Yes | Reference |  | Reference |  |
| No | 1.3 (1.23-1.39) | <0.001 | 1.1 (1.05-1.21) | **<0.001** |
| **On a dietary plan recommended by healthcare worker** |  |  |  |  |
| Yes | Reference |  | Reference |  |
| No | 1.3 (1.19-1.45) | <0.001 | 1.1 (1.02-1.22) | **0.003** |
| **Belief that regimen complexity impairs adherence** |  |  |  |  |
| No | Reference |  | Reference |  |
| Yes | 1.4 (1.28-1.48) | <0.001 | 1.2 (1.12-1.34) | **<0.001** |
| **Perceived impact of stress on glycemia** |  |  |  |  |
| No | Reference |  | Reference |  |
| Yes | 1.4 (1.29-1.58) | <0.001 | 1.1 (1.08-1.28) | **<0.001** |
| **Access to peer support groups** |  |  |  |  |
| Yes | Reference |  | Reference |  |
| No | 1.4 (1.33-1.53) | <0.001 | 1.2 (1.08-1.32) | **<0.001** |
| **Costs in accessing care** |  |  |  |  |
| No | Reference |  | Reference |  |
| Yes | 1.5 (1.38-1.60) | <0.001 | 1.2 (1.17-1.33) | **<0.001** |
| **Receipt of comprehensive diabetes care package** |  |  |  |  |
| Yes | Reference |  | Reference |  |
| No | 1.2 (1.13-1.27) | <0.001 | 1.0 (0.99-1.06) | 0.239 |

## Discussion

This study assessed predictors of poor glycemic control among adults attending a peri-urban diabetic clinic in Wakiso district, Uganda using modified Poisson regression analysis. Overall, 67.8% of the participants had poor glycemic control, indicating a substantial burden of uncontrolled diabetes among patients attending Kasangati Health Centre IV. This prevalence is comparable to findings from other studies conducted in sub-Saharan Africa, where poor glycemic control among patients with diabetes has ranged between 60% and 80% [13, 14].

The high prevalence of poor glycemic control observed in this study may reflect the combined influence of socioeconomic barriers, limited continuity of chronic care, inadequate lifestyle modification support, psychosocial challenges, and rising treatment burden among patients with diabetes in low-resource settings. In peri-urban communities such as Wakiso district, rapid urbanization, changing dietary patterns, sedentary lifestyles, and increasing out-of-pocket healthcare expenditures may further complicate diabetes self-management and long-term glycemic control. In addition, health systems in many low- and middle-income countries remain largely oriented toward acute care delivery, which may limit the effectiveness of long-term chronic disease management services such as regular monitoring, adherence counseling, and psychosocial support for patients with diabetes [15].

Older age was independently associated with poor glycemic control. Participants aged 40–64 years and those aged ≥65 years were more likely to experience poor glycemic control compared to younger participants. This finding may be attributed to progressive pancreatic beta-cell dysfunction, longer duration of disease, increasing treatment burden, and the presence of age-related comorbidities that complicate diabetes management. Older adults may also experience challenges related to reduced physical activity, cognitive decline, and adherence difficulties associated with multiple medications. Similar findings have been reported in studies conducted in Ethiopia and Nigeria, where older age groups were significantly associated with suboptimal glycemic outcomes [16, 17].

Low income level was a strong predictor of poor glycemic control. Participants with low income were significantly more likely to have uncontrolled blood glucose levels compared to those with higher income. Socioeconomic disadvantage can negatively affect diabetes management through inability to afford medications, transportation to health facilities, recommended diabetic diets, and routine laboratory investigations. Financial hardship may also reduce access to health information and supportive diabetes care services. Similar associations between low socioeconomic status and poor glycemic control have been reported in studies from low- and middle-income countries [18]. This finding highlights the broader structural inequalities that influence chronic disease outcomes beyond individual patient behaviors alone.

Polypharmacy was significantly associated with poor glycemic control. Participants taking two or more concurrent medications were more likely to have uncontrolled glycemia compared to those taking fewer medications. Multiple medications may increase treatment complexity, polypharmacy, medication fatigue, and forgetfulness, ultimately affecting adherence. In addition, patients requiring multiple medications are often those with more advanced disease or multiple comorbid conditions, which further complicate glycemic management. Similar findings have been reported in studies among patients with diabetes in resource-limited settings where increasing medication burden was associated with reduced adherence and poor treatment outcomes [18, 19, 20].

Non-adherence to regular clinic follow-up was also identified as a significant predictor of poor glycemic control. Patients who failed to attend their routine clinical follow-ups had poor glycemic outcomes as compared to those who attended their clinical follow-ups. Regular follow-up is essential for monitoring glycemic trends, adjusting treatment regimens, reinforcing adherence counseling, and identifying diabetes-related complications early. In other words, missing scheduled clinic appointments could result in delayed treatment optimization, poor continuity of care, and inadequate self-management support. Similar findings have been reported in previous studies conducted in Ethiopia and Kenya where non-attendance of clinic follow-ups was found to predict poor glycemic outcomes among patients with diabetes mellitus [21, 22].

Medication side effects were independently associated with poor glycemic control. Participants who experienced side effects were more likely to have uncontrolled glycemia compared to those without side effects. Medication-related adverse effects may discourage continued medication use, contribute to irregular dosing practices, or reduce adherence due to fear of worsening symptoms. This finding underscores the importance of routine assessment, counseling, and management of adverse drug effects during diabetes care. Similar findings have been documented in studies examining treatment adherence among diabetic patients [23, 24].

Lifestyle-related factors including physical inactivity and failure to follow recommended dietary plans were significantly associated with poor glycemic control. Participants who did not engage in regular physical exercise and those not adhering to dietary recommendations had increased likelihood of uncontrolled glycemia. Physical activity improves insulin sensitivity and glucose utilization, while healthy dietary practices remain central to diabetes management. Failure to sustain these lifestyle modifications may therefore contribute to persistent hyperglycemia. Similar findings have been reported in previous studies where sedentary behavior and poor dietary adherence were strongly linked to poor glycemic outcomes [25, 26]. These findings reinforce the importance of strengthening lifestyle counseling within routine diabetes care services.

Factors related to psychosocial wellbeing and the treatment process also had a considerable impact on glycemic status. Participants who believed that treatment regimen complexity impaired adherence were more likely to experience poor glycemic control. Complex treatment regimens may increase pill burden, dosing confusion, treatment fatigue, and forgetfulness, particularly among patients with limited health literacy or inadequate counseling support. In low-resource settings, regimen complexity may further worsen adherence where patients already face financial barriers, inconsistent medication access, and limited continuity of care. Poor adherence resulting from complex medication schedules may consequently contribute to persistent hyperglycemia and suboptimal glycemic outcomes. Similar findings have been reported in studies linking medication regimen complexity, poor adherence, and uncontrolled glycemia among patients with type 2 diabetes [27, 28].

Similarly, participants who perceived stress as affecting their glycemia had increased likelihood of poor glycemic control. Psychological stress may negatively affect glycemic outcomes through both physiological where stress-related hormonal responses involving cortisol and catecholamine release may contribute to elevated blood glucose levels where and behavioral where stress may also reduce adherence to medications, disrupt dietary practices, impair sleep, and limit engagement in physical activity. In low-resource settings, psychosocial stress may be further compounded by financial insecurity, chronic medication costs, and long-term disease burden, which may collectively undermine diabetes self-management practices. This finding highlights the importance of integrating mental health screening, psychosocial counseling, and patient support interventions into routine diabetes care services. Similar associations between psychosocial stress, diabetes distress, and poor glycemic outcomes have been reported in previous studies among patients with diabetes [29, 30].

Lack of access to peer support groups was independently associated with poor glycemic control. These groups are likely to benefit those with diabetes by providing emotional support, common experiences, education, motivation, and reinforcement of self-management skills. In settings where resources are inadequate, peer networks might provide additional help with dealing with the stigma, frustration, and stress associated with their condition, as well as limited access to counseling. Patients without such support systems may therefore experience reduced adherence, poorer coping mechanisms, and suboptimal glycemic outcomes. Similar findings have been reported in a studies from Uganda demonstrating that social support and peer-based interventions can improve diabetes self-management and treatment adherence among patients with chronic disease [31].

Financial barriers to accessing diabetes care were significantly associated with poor glycemic control. Participants who experienced costs related to transportation, medications, laboratory investigations, and clinic attendance were more likely to have uncontrolled glycemia. Unlike acute illnesses, diabetes requires lifelong treatment, regular monitoring, continuous medication use, and sustained lifestyle modification, all of which may impose substantial financial burden on patients and households in low-resource settings. In peri-urban communities where out-of-pocket health expenditures remain high, financial constraints may compromise treatment adherence, continuity of care, and timely access to monitoring services. This finding highlights the broader influence of socioeconomic inequalities on chronic disease outcomes and underscores the need for policies that improve affordability and accessibility of diabetes care services. Similar findings have been reported in studies from sub-Saharan Africa where healthcare costs negatively affected diabetes treatment adherence and glycemic outcomes [32, and 33].

## Study limitations

The cross-sectional study design limits the ability to establish causal relationships between the identified predictors and poor glycemic control. Second, some variables such as adherence to exercise, dietary practices, stress, and clinic follow-up were self-reported and may therefore have been affected by recall bias or social desirability bias. Third, the study was conducted at a single peri-urban health facility, which may limit the generalizability of the findings to other diabetic populations in Uganda, particularly rural or tertiary healthcare settings. Additionally, some potentially important factors such as detailed dietary intake, depression, and health literacy were not comprehensively assessed. Despite these limitations, the study provides important evidence on the burden and predictors of poor glycemic control among adults attending a peri-urban diabetic clinic in Uganda.

## Conclusion and recommendations

Poor glycemic control among adults attending the diabetic clinic at Kasangati Health Centre IV was high. Various independent predictors of poor glycemic control were identified. These included older age, low income level, polypharmacy, non-adherence to regular follow-up schedules, medication side effects, physical inactivity, failure to follow recommended dietary plans, perceived treatment regimen complexity, stress affecting glycemia, lack of peer support groups, and costs involved in accessing diabetes care.

These findings highlight that poor glycemic control is driven by a combination of socioeconomic, behavioral, psychosocial, clinical, and health-system-related factors. Therefore, there is need a for comprehensive patient-centered and multi-sectoral interventions aimed at strengthening diabetes education, improving adherence to treatment and follow-up schedules, promoting healthy dietary and physical activity practices, integrating psychosocial support into routine diabetes care, establishing peer support groups, and reducing the financial burden associated with diabetes management. In addition, the Ministry of Health and district health authorities should strengthen diabetes care services at lower-level health facilities through ensuring availability of diabetes medications, HbA1c testing supplies, and adequately trained healthcare workers. Further longitudinal and multicenter studies should also be conducted to establish causal relationships between the identified predictors and poor glycemic control and to explore the role of mental health, family support, health literacy, and healthcare system factors in diabetes management outcomes in Uganda.

## Declarations

### Availability of data and materials

All raw data required to replicate the results of this study are included within the Supporting Information files (S1 File).

### Competing interests

The authors have declared that no competing interests exist.

### Consent for publication

Not applicable.

### Funding

The authors received no specific funding for this work.

### Author contributions

L.N.Y.K and R.K conceived the study, designed the research, collected the data, and drafted the manuscript. R.K analyzed the data, developed the visualizations, and wrote the results section of the manuscript. D.M, E.R, O.N, M.W.W.D, and P.S supervised the study, provided critical revisions, and offered expert guidance throughout the study design, data interpretation, and manuscript preparation. All authors read and approved the final manuscript for submission.

## Acknowledgements

We acknowledge the Kasangati Health Centre IV administrative authorities for granting permission to conduct the study.

## Supporting Information

### S1 File. Dataset used for analysis

An anonymized dataset containing variables used in the quantitative analysis of the factors associated with poor glycemic control at Kasangati Health Centre IV.

## List of abbreviations and acronyms

aPR: Adjusted Prevalence Ratio
CI: Confidence Interval
cPR: Crude Prevalence Ratio
DM: Diabetes Mellitus
HbA1c: Glycated Hemoglobin
REC: Research and Ethics Committee
SPSS: Statistical Package for Social Sciences
WHO: World Health Organization

## Notes

### Competing Interest Statement

The authors have declared no competing interest.

### Author Declarations

This study was reviewed and approved by the Makerere University, School of Public Health Research and Ethics Committee under reference number MakSPH-REC_502.

